# Prognostic factors of chest CT findings for ICU admission and mortality in patients with COVID-19 pneumonia

**DOI:** 10.1101/2020.10.30.20223024

**Authors:** Mohammad Ali Kazemi, Hossein Ghanaati, Behnaz Moradi, Mohammadreza Chavoshi, Hassan Hashemi, Samira Hemmati, Pouria Rouzrokh, Masoumeh Gity, Zahra Ahmadinejad, Hamed Abdollahi

**Author notes:** **Corresponding author:** Mohammadreza chavoshi, MD, Shariati hospital, Ale Ahmad highway, Tehran, Iran. Competing Interest Statement: The authors have declared no competing interest.

## Abstract

**Background:** Studies have shown that CT could be valuable for prognostic issues in COVID-19

**Objective:** to investigate the prognostic factors of early chest CT findings in COVID-19 patients.

**Materials and Methods:** This retrospective study included 91 patients (34 women, and 57 men) of RT-PCR positive COVID-19 from 3 hospitals in Iran between February 25, 2020, to march 15, 2020. Patients were divided into two groups as good prognosis, discharged from the hospital and alive without symptoms (48 patients), and poor prognosis, died or needed ICU care (43 patients). The first CT images of both groups that were obtained during the first 8 days of the disease presentation were evaluated considering the pattern, distribution, and underlying disease. The total CT-score was calculated for each patient. Univariate and multivariate analysis with IBM SPSS Statistics v.26 was used to find the prognostic factors.

**Results:** There was a significant correlation between poor prognosis and older ages, dyspnea, presence of comorbidities, especially cardiovascular and pulmonary. Considering CT features, peripheral and diffuse distribution, anterior and paracardiac involvement, crazy paving pattern, and pleural effusion were correlated with poor prognosis. There was a correlation between total CT-score and prognosis and an 11.5 score was suggested as a cut-off with 67.4% sensitivity and 68.7% specificity in differentiation of poor prognosis patients (patients who needed ICU admission or died. Multivariate analysis revealed that a model consisting of age, male gender, underlying comorbidity, diffused lesions, total CT-score, and dyspnea would predict the prognosis better.

**Conclusion:** Total chest CT-score and chest CT features can be used as prognostic factors in COVID-19 patients. A multidisciplinary approach would be more accurate in predicting the prognosis.

## Introduction

After 2 outbreaks of coronavirus by SARS-COV and the Middle East Respiratory Syndrome Coronavirus in 2003 and 2012 respectively, the world faced the 3^rd^ outbreak since December 2019 [1-4]. Although the mortality rate is lower than 2 previous outbreaks [5], the striking speed of disease spread lead to more death worldwide.

The real-time reverse transcriptase-polymerase chain reaction (RT-PCR) test has been proposed as a specific test for COVID-19 but the sensitivity is as low as 60-70% [6,7]. Given the limited access for test kits in many countries, there is a high demand for additional diagnostic tests. It is suggested that chest CT scan may have higher sensitivity than RT-PCR for diagnosis of COVID-19 [6,7]. CT scan may help to diagnose the disease in clinically suspicious cases with primary RT-PCR negative results [8,9]. According to the stage of the disease, CT scan shows different findings [10,11]. The main findings are ground-glass opacity (GGO), consolidation or both in peripheral and lower zones of both lungs [12]. However, additional findings like crazy-paving pattern, multilobar involvement, and increasing lung consolidations are seen as the disease progresses [7]. Some of these findings can determine the severity and extension of the disease [13,14].

Although some laboratory and clinical parameters have been proposed to evaluate the severity of the disease [15-17], limited data is available about the correlation of CT scan findings and disease prognosis. Although CT scan is more useful in the detection or exclusion of viral pneumonia including COVID-19, some CT scan findings may help to investigate the severity of disease and determine the therapeutic approach for the patient.

### Objective

This study aimed to investigate what findings on CT scan can predict mortality or the need for ICU admission in laboratory definite cases of COVID-19.

## Method and material

This study was approved by the Ethics committee of Tehran University of Medical Sciences with the ethical code of IR.TUMS.MEDICINE.REC.1399.095 and written informed consent was waived.

### Patients

The data of the patients from 3 medical centers were evaluated from February 25, 2020, to April 25, 2020, retrospectively. The study population was defined as patients older than 18 years old and with definite RT-PCR positive diagnosis for COVID-19 who had undergone CT scan examination in less than 8 days from the beginning of their symptoms. If the time interval between the RT-PCR assay and a chest CT scan was more than 7 days, the patient was excluded at the initial evaluation.

We enrolled 104 patients in the study: 28 patients who had expired (Group A), 16 Patients with ICU admission and subsequent recovery (Group B), and 60 patients with no need for ICU admission or outpatient treatment (Group C). Follow up evaluation was done by telephone interview with all the discharged patients and their health status were checked by a medical doctor. All of the discharged patients were alive until May 20, 2020. 12 patients from group C had missing contact data and were excluded due to loss of follow up. Another case from group A was also excluded as a synchronous massive pulmonary thromboembolism was found on her CT scan that could have led to her death. Finally, the total number of 91 patients entered the study. Clinical symptoms and history of previous diseases were documented for all patients. Comorbidities were evaluated as cardiovascular disease, pulmonary disease, and other disease. diabetes mellitus and hypertension cases were included in cardiovascular group if history of cardiac complication was positive. Smoking history was asked from the patients and if related pulmonary complications were occurred the history was considered positive.

### CT scan protocol

Non-enhanced chest CT scan images were obtained in the supine position, using CT scan systems (SOMATOM Emotion 16 scanner; Siemens). To minimize motion artifacts, CT scan images were acquired during a single inspiratory breath-hold. For minimizing patient radiation exposure, we used following acquisition parameters: tube voltage = 80-110 kVp, effective current 60-80 mA, pitch = 1-1.5, matrix = 512 × 512, slice thickness = 5 mm (reconstructed slice thickness= 1.5 mm), and pulmonary U90S kernel. The reconstructed images were sent to the picture archiving and communication system (PACS). The low dose CT scan protocol was recommended by the Iranian Society of Radiology COVID-19 Consultant Group (ISRCC) and didn’t make any problem in image interpretations [18].

### Imaging interpretation

All chest CT scans were reviewed by two radiologists concurrently with both lung (width, 1500 HU; level,-700 HU) and mediastinal (width, 350 HU; level, 40 HU) windows and after a final agreement, the prepared check-list was filled. During the review, both radiologists were blinded about the patent’s information and outcome. The CT scan evaluation was performed in 5 major fields: morphology, distribution, CT scan involvement score, associated pulmonary lesions, and mediastinal findings. Morphology features included ground-glass opacity GGO, consolidation, mixed type with >50% GGO, mixed type with >50% consolidation, crazy-paving pattern (a combination of GGO with superimposed interlobular and intralobular septal thickening), reversed halo sign, nodular pattern, intralesional bronchial distortion, and linear opacity. Four main distribution patterns were evaluated: peripheral (pleural based or/and pleural sparing), central (Patchy opacities that extended to the lung hila and showed lobar bronchial contact without obvious peri-broncho-vascular appearance), peri-broncho-vascular (involvement with the disease and not edema) and diffuse (randomly central and peripheral lesions). We expected edema as fine smooth interstitial peribronchovascular thickening usually with other imaging supporting findings. Also, the involvement of anterior areas (the involvement of anterior one-fourth of lung periphery in both upper and lower areas), and paracardiac areas (more than 2 cm in contact with pericardium at lingula and right middle lobe) were evaluated. All lung lobes were visually evaluated for CT scan involvement scores. Each lobe received 0 (non-involvement), 1 (less than 5% involvement), 2 (5-25% involvement), 3 (26-49% involvement), 4 (50-75% involvement) and, 5 (>75% involvements) [13]. A total CT-score was recorded with a range of 0-25. Furthermore, a mean lung score was calculated for each lung by dividing the sum of individual lobar scores of each lung by 3 in the right and 2 in the left lungs. Underlying pulmonary disease as bronchiectasis, emphysema, fibrosis or mass was recorded. The mediastinal setting was assessed for lymphadenopathy, pulmonary artery enlargement, pleural effusion, and pericardial effusion.

### Statistical analysis

IBM SPSS Statistics v.25 (Armonk, NY: IBM Corp) was used for statistical analysis. To determine the prognosis of the patients, groups A and B from above were considered together as the “poor-prognosis” group and group C was considered as the “good-prognosis” group. Frequencies and descriptive statistics were then calculated for all variables. To determine the relationship between independent (all demographic, clinical and imaging features) and dependent (prognosis) variables, Pearson’s chi-square test and the Mann-Whitney U test were used where appropriate. The relationship between the total CT-score and the prognosis was determined by both univariate and multivariate binary logistic regression (non-stepwise) tests. For the latter, a combination of other clinically and statistically significant variables was also considered as inputs along with the total CT-score. Receiver operating characteristic (ROC) curves were plotted for both models and cut-offs with the best trade-off for sensitivity and specificity were proposed to predict the output. The Spearman rank-order correlation test was used to determine the relationship between the total CT-score and the number of days it took patients from the beginning of the symptoms to the day of doing the CT scan (hereafter called the CT scan day). The relationship between the mean CT-score of each lung and the ipsilateral presence of the pleural effusion was assessed using the Man-Whitney-U test. P values less than.05 were considered significant.

## Results

Ninety-one [57 (62.6%) men and 34 (37.4%) women] patients were included in the study. The average age of patients was 58.04 years (SD=16.5) and they were admitted in the hospital for a variation of 1 to 18 days (mean=5.90, SD=4.21). 48 (52.8%) patients did not need ICU care and survived, 16 (17.6%) were admitted to the ICU and survived, 11 (12.0%) were admitted to the ICU and died and 16 (17.6%) died without being admitted to the ICU. Overall, the “good-prognosis” group consisted of 48 (52.7%) and the “poor-prognosis” group consisted of 43 cases (47.3%). Only 8 (8.8%) cases had normal CT scans and 83 (91.2%) cases had either ground-glass opacity, consolidations or both (figure 1). Among the clinical findings, the most common was fever with 81 (89.0%) of affected cases. 42 (46.2%) of patients had no comorbidities while 18 (19.8%) had cardiovascular, 11 (12.1%) had pulmonary, 8 (8.8%) had diabetes mellitus (DM) and 12 (13.2%) had other comorbidities. CT scan and clinical findings/conditions are summarized in Table 1 and 2.

**Table 1.** Frequencies of Clinical findings

| Finding | Frequency |
| --- | --- |
| Fever | 81 (89.0%) |
| Cough | 67 (73.6%) |
| GI symptoms | 2 (2.2%) |
| Dyspnea | 40 (44.0%) |
| Underlying comorbidity | Cardiac: 18 (19.8%)<br>Pulmonary: 11(12.1%)<br>Diabetes Mellitus: 8 (8.8%)<br>Others: 12 (13.2%) |
| ICU Admission | 27 (29.7%) |
| Death | 27 (29.7%) |
| Prognosis | Good: 48 (52.7%)<br>Bad: 43 (47.3%) |

Good prognosis: no need for ICU care and discharged, Bad prognosis: dead or need for ICU care

**Table 2:** Frequencies of CT scan findings

| CT-scan findings | Frequencies |
| --- | --- |
| **Main CT findings** |  |
| Normal CT scan | 8 (8.8%) |
| Ground glass opacities / Consolidation | Pure GGO: 10 (11.0%)<br>GGO >50%: 43 (47.3%)<br>Pure Consolidation: 3 (3.3%)<br>Consolidation > 50%: 24 (26.4%) |
| Crazy paving | 7 (7.7%) |
| Reversed halo sign | 21 (23.1%) |
| Nodular pattern | 2 (2.2%) |
| Intralesional bronchial distortion | 30 (33.0%) |
| Linear opacities | 44 (48.3%) |
| **Lesion distribution** |  |
| Peripheral distribution | Pleural_based: 19 (20.9%)<br>Pleural_sparing: 11 (12.1%)<br>Pleural_based and pleural sparing: 53 (58.2%) |
| Central distribution | 52 (57.15%) |
| Peribronchovascular distribution | 20 (22.0%) |
| Diffuse distribution | 12 (13.2%) |
| Anterior distribution | 60 (65.9%) |
| Paracardiac distribution | 30 (33.0%) |
| **Additional findings** |  |
| Associated bronchiectasis | 1 (1.1%) |
| Associated emphysema | 4 (4.4%) |
| Associated fibrosis | 0 |
| Associated mass | 0 |
| Lymphadenopathy | 9 (9.9%) |
| pulmonary artery enlargement (>30 mm) | 14 (15.4%) |
| Pleural effusion | Trace:49 (53.8%)<br>Mild: 8 (8.8%) |
| Side of pleural effusion | Right: 21 (23.1%)<br>Left: 3 (3.3%)<br>Bilateral: 33 (36.3%) |

**Figure 1.**
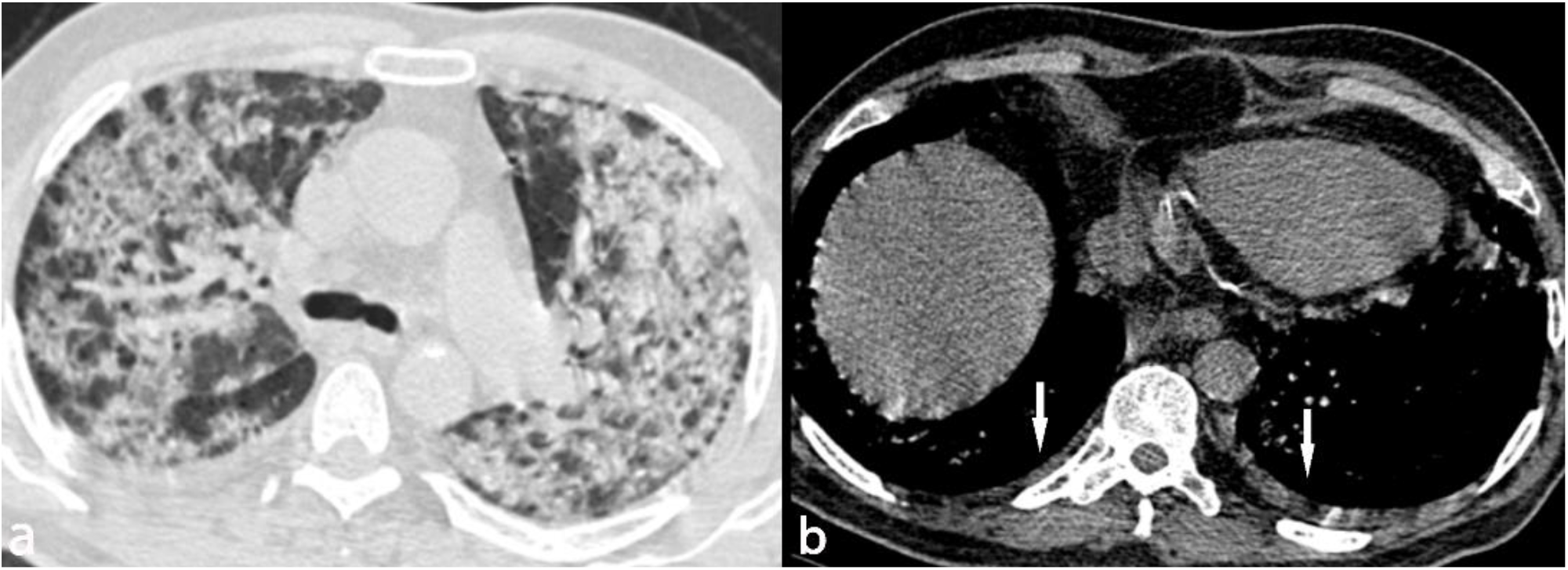
Image in a 68-years-old man who presented with dyspnea from 4 days ago. (a) Axial thin286 section unenhanced CT scan shows bilateral diffuse consolidation and ground glass opacities. (b) Axial thin-section unenhanced CT scan shows bilateral mild pleural effusion. The patient was admitted to the ICU 10 days after this CT scan.

An increase in age and the male gender was associated with poor prognosis. Having underlying comorbidity was also associated with poor-prognosis; 72% of cases with cardiovascular, 90.9% of cases with pulmonary, 50% of cases with DM, and 41.7% of cases with other comorbidities had poor-prognosis. No significant association was detected between having fever, cough or diarrhea and the prognosis, yet dyspnea was associated with poor prognosis. Among CT scan findings, the crazy-paving pattern, peripheral distribution, diffuse distribution, anterior distribution, paracardiac distribution, lymphadenopathy, main pulmonary artery dilation (above 30mm) and pleural effusion were associated with poor prognosis (figures 2-5). On the other hand, the reversed halo sign was associated with a better prognosis. Test types and the test results of statistically significant associations between patient prognosis and demographic, clinical and CT scan findings are summarized in Table 3. No case of pericardial effusion was seen in this study.

**Table 3:** Test types and test results of statistically significant associations between patient prognosis and demographic, clinical and CT scan findings

| **Independent variable** | **Statistical test used** | **Test results** | **Good-prognosis** | **Poor-prognosis** |
| --- | --- | --- | --- | --- |
| Age | Man-Whitney U | U=410, p<.001 | Median: 51 | Median: 68 |
| Gender | Chi-square | X<sup>2</sup>(1)=4.83, p=.028,<br>Phi=0.23 | Male: 43.9%<br>Female:67.6% | Male: 56.1%<br>Female: 32.4% |
| Underlying comorbidity | Chi-square | X<sup>2</sup>(4)=20.56, p<.001,<br>Phi = 0.47 | Cardiac: 28.0%<br>Pulmonary: 9.1%<br>DM: 50.0%<br>Others: 58.3% | Cardiac: 72.0%<br>Pulmonary: 90.9%<br>DM: 50.0%<br>Others: 41.7% |
| Hospital admit duration (days) | Man-Whitney U | U=793, p=.056 | Median: 4 | Median: 6 |
| Dyspnea | Chi-square | X<sup>2</sup>(1)=11.73, p=.001;<br>Phi=.359 | No: 68.6%<br>Yes: 32.5% | No: 31.4%<br>Yes: 67.5% |
| Crazy-paving | Chi-square | X<sup>2</sup>(1) = 4.50, p=.034,<br>Phi = 0.22 | No: 56.0%<br>Yes: 14.3% | No: 44.0<br>Yes: 85.7 |
| Reverse halo sign (RHS) | Chi-square | X<sup>2</sup>(1)=3.82, p=.051,<br>Phi=-0.20 | No: 47.1%<br>Yes: 71.4% | No: 52.9<br>Yes: 28.6 |
| Peripheral distribution | Chi-square | $X^2(1)=7.8$, $p=.005$,<br>Phi=0.29 | No: 100%<br>Yes: 48.2% | No: 0%<br>Yes: 51.8% |
| Diffuse distribution | Chi-square | $X^2(1)=10.94$, $p=.001$,<br>Phi=.34 | No: 59.5%<br>Yes: 8.3% | No: 40.5%<br>Yes: 91.7% |
| Anterior distribution | Chi-square | $X^2(1)=6.26$, $p=.01$,<br>Phi=0.26 | No: 71%<br>Yes: 43.3% | No: 29.0%<br>Yes: 56.7% |
| Paracardiac distribution | Chi-square | $X^2(1)=6.76$, $p=.009$,<br>Phi=0.27 | No: 62.3%<br>Yes: 33.3% | No: 37.7%<br>Yes: 66.7% |
| Lymphadenopathy | Chi-square | $X^2(1)=3.88$ $p=.049$,<br>Phi=0.20 | No: 56.1%<br>Yes: 22.2% | No: 43.9%<br>Yes: 77.8 |
| Pulmonary artery dilation | Chi-square | $X^2(1)=3.73$ $p=.053$,<br>Phi=0.20 | No: 57.1%<br>Yes: 28.6% | No: 42.9%<br>Yes: 71.4% |
| Pleural effusion | Chi-square | $X^2(1)=10.20$, $p=.006$,<br>Phi=0.33 | No: 70.6%<br>Trace: 46.9%<br>Mild: 12.5% | No: 29.4%<br>Trace: 53.1%<br>Mild: 87.5% |

**Figure 2.**
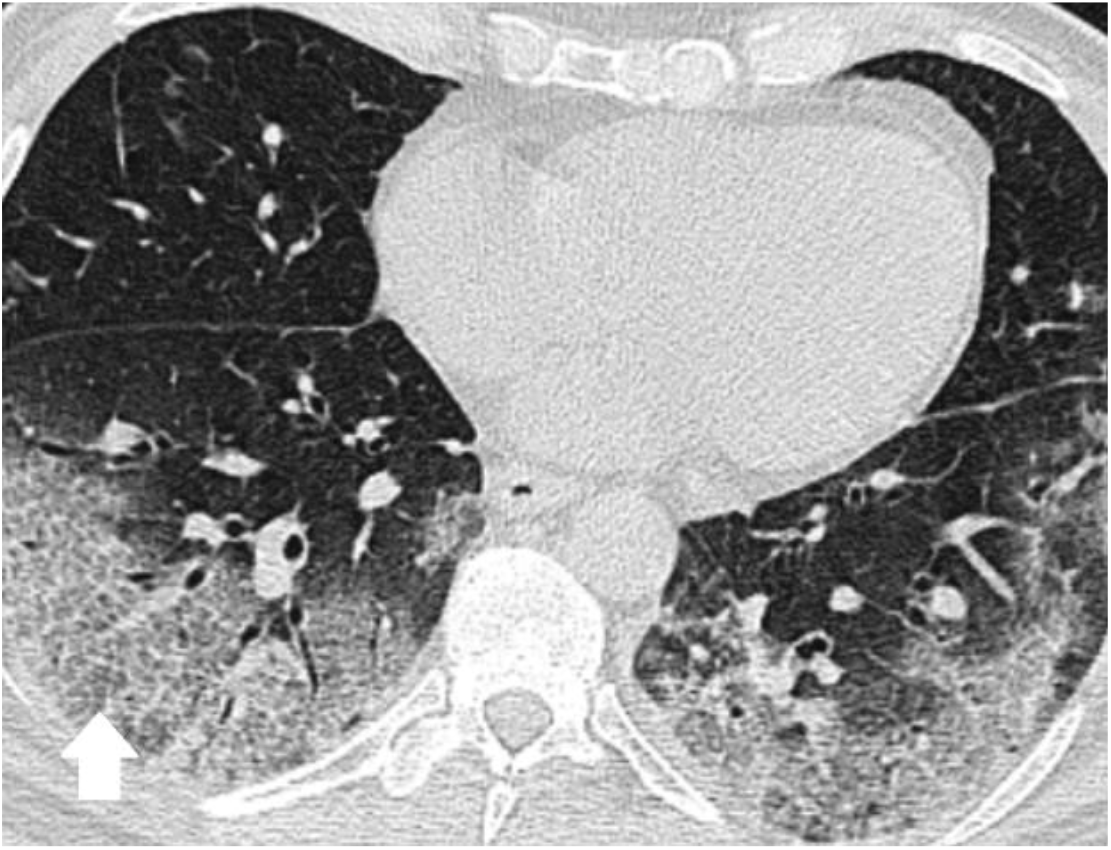
Image in a 55-year-old man who presented with fever and cough from 8 days ago. Axial thin section unenhanced CT scan shows peripheral opacities with crazy paving appearance more prominent on right side. The patient was admitted to the ICU 4 days after this CT scan.

**Figure 3.**
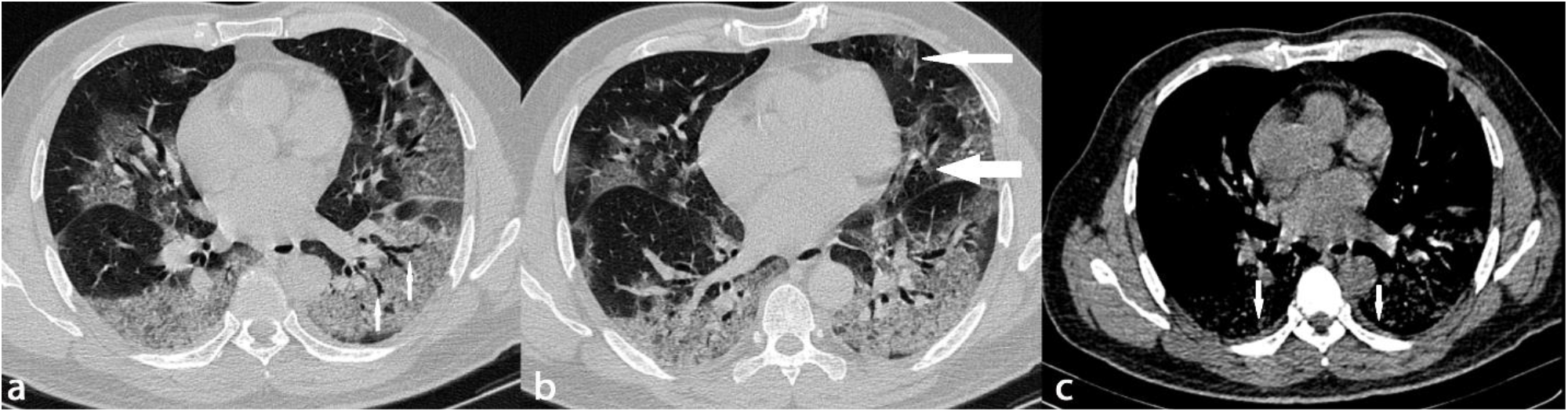
A 59-year-old man who presented with cough, fever and dyspnea from 7 days ago. (a) Axial thin section unenhanced CT scan shows bilateral peripheral GGO with intralesional bronchial distortion. (b) Axial thin-section unenhanced CT scan with paracardiac (wide arrow) and anterior (narrow arrow) extension of the opacities. This density gradient raises concerns about ARDS formation (c) Axial thin section unenhanced CT scan shows bilateral trace pleural effusion. The patient was admitted to the ICU one day later.

**Figure 4.**
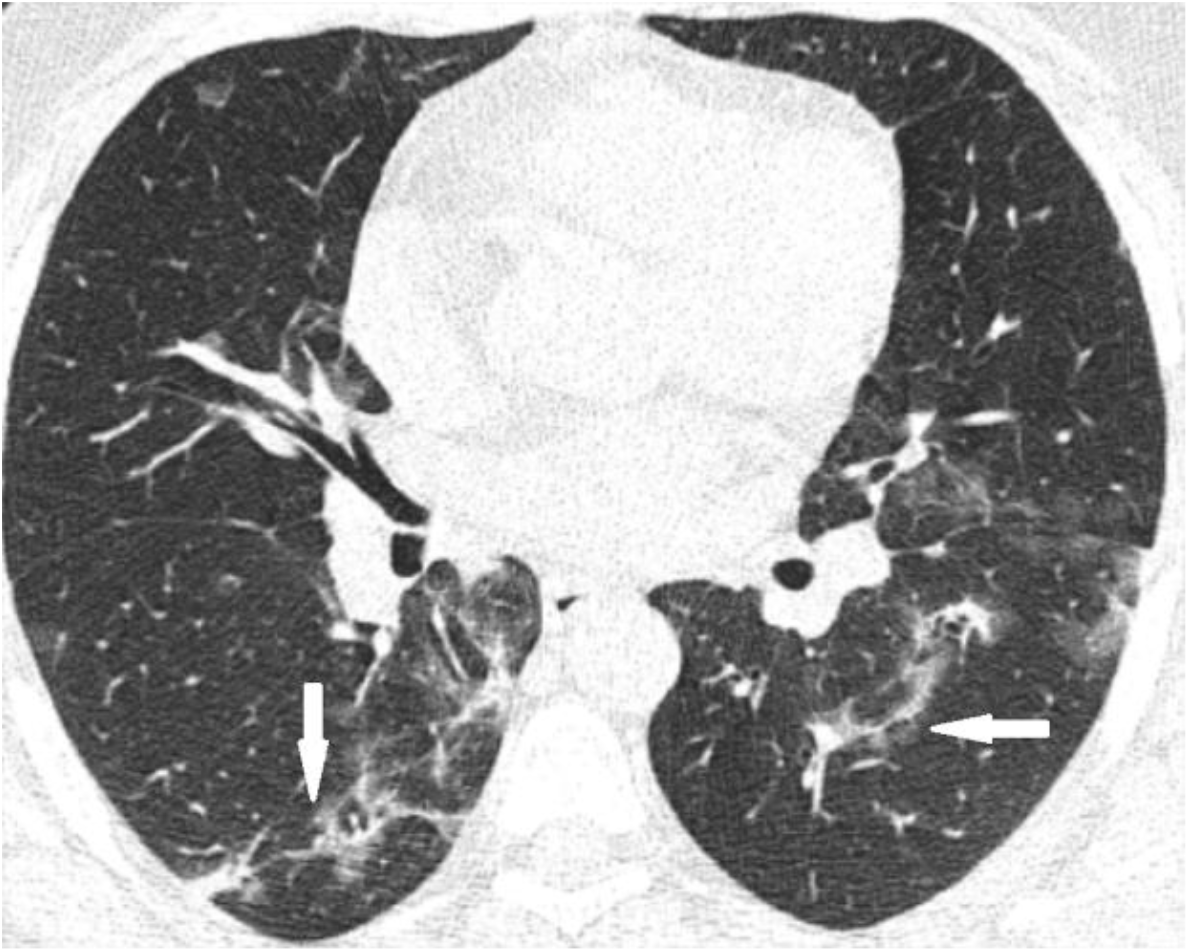
Image in a 40-year-old man who presented with history of fever and cough from 7 days ago. Axial thin-section unenhanced CT scan shows bilateral peripheral linear opacities. The patient was treated on an outpatient basis.

**Figure 5:**
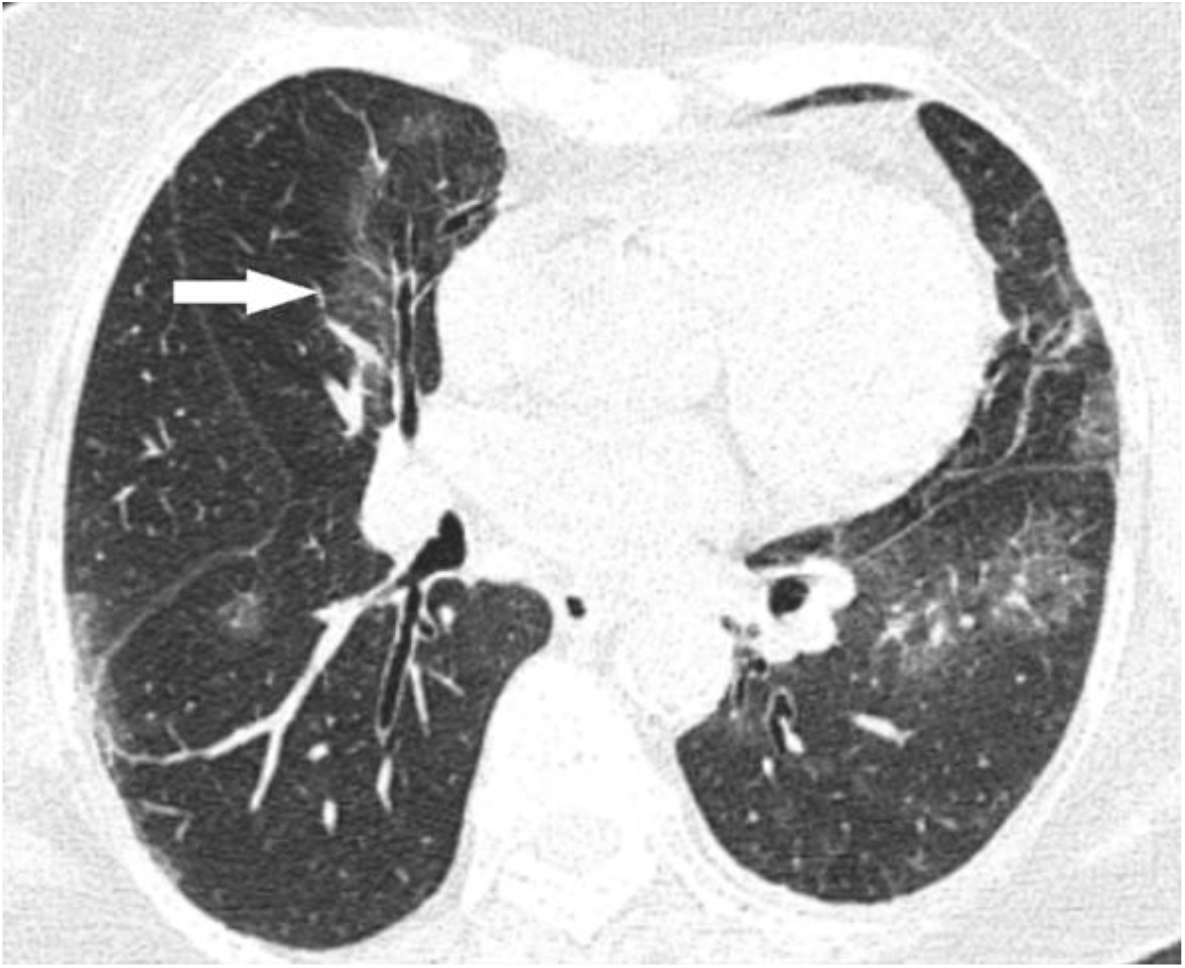
A 87-years-old woman who presented with history of cough and dyspnea from 6 days before. Axial thin-section unenhanced CT scan shows peripheral ground glass opacities with paracardiac and anterior extension. Unfortunately, the patient died 12 days after this exam.

The univariate binary logistic regression model was able to predict the prognosis by the total CT-score (X^2^(1) = 15.02, p<.001). The model explained 21.8% (Nagelkerke R2) of the variance in prognosis and correctly predicted 68.1% of cases. Increasing the total CT-score was associated with an increase in the likelihood of poor prognosis (OR=1.17). The ROC curve for this univariate model is plotted (figure 6). The AUC for the model is 0.73 (p<.001) and by choosing a cut-off of 11.5 for the total CT-score, the resulting sensitivity and specificity would be 67.4% and 68.7% respectively. A multivariate binary logistic regression model was also developed including total CT-score, male gender, age, underlying pulmonary or cardiovascular comorbidity, dyspnea, and diffuse distribution of CT scan findings as input variables (X^2^(8)=21.65, p=.006). Table 4 shows the statistical significance and odds ratios for each variable in the latter model. Except for underlying comorbidity, all other input variables significantly contribute to the model. The model explained 64.4% (Nagelkerke R2) of the variance in prognosis and correctly predicted 86.8% of cases. The ROC curve for this univariate model is plotted (figure 7). The AUC for the model is 0.94 (p<.001) and by choosing a cut-off of 0.61 for the model output, the resulting sensitivity and specificity would be 81.4% and 79.0% respectively.

**Table 4:** Statistical significance and odds ratios for independent variables of a multivariate binary logistic model to predict patients’ prognosis.

| **Independent variable** | **Significance** | **Odds Ratio** | **95% confidence interval lower** | **95% confidence interval upper** |
| --- | --- | --- | --- | --- |
| Total CT-score | .050 | 1.17 | 1.00 | 1.38 |
| Male gender | .004 | 0.08 | 0.01 | 0.46 |
| Age | .003 | 1.08 | 1.02 | 1.14 |
| Comorbidity (cardiovascular and pulmonary) | .177 | 2.93 | 0.61 | 13.99 |
| Dyspnea | .038 | 3.90 | 1.07 | 14.16 |
| Diffuse distribution | .038 | 17.27 | 1.17 | 254.30 |
| Constant | .001 | 0.001 | .. | .. |

**Figure 6:**
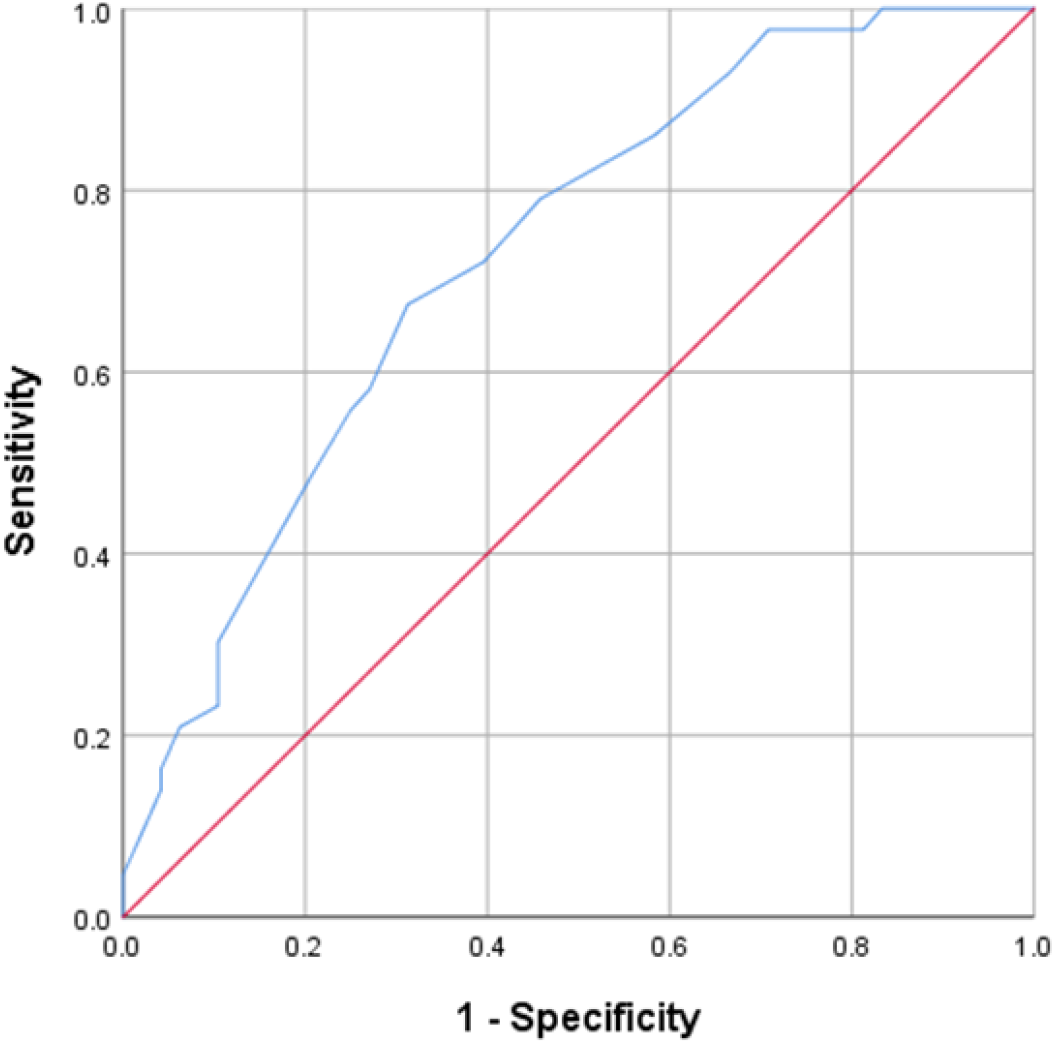
ROC Curve for a univariate model to predict patients’ prognosis based on their Total CT-score.

**Figure 7:**
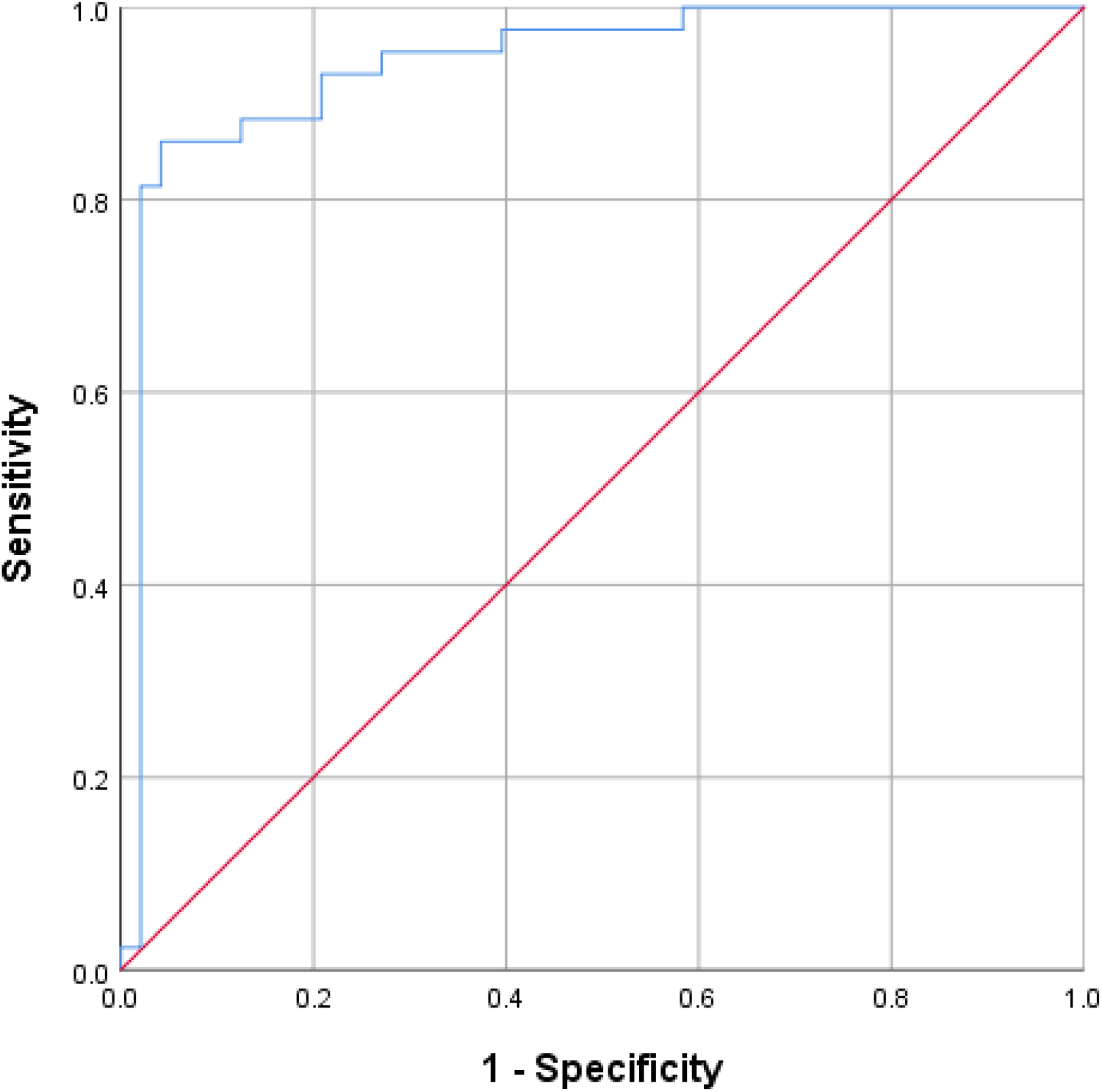
ROC Curve for a multivariate model to predict patients’ prognosis based on Total CT-score, gender, age, underlying pulmonary or cardiac comorbidity, dyspnea, and diffuse distribution of CT scan findings.

An increase in the total CT-score had a weak correlation with an increased CT scan day (Spearman’s rank two-tailed Correlation, rho=.26, p=.023), indicating that as the disease progresses in the first week of symptoms, pulmonary involvement in CT becomes more severe. Also, the mean of CT-score for lobes of each lung (mean CT-score) was statistically different between the patients with or without ipsilateral pleural effusion (Man-Whitney U test, p<0.001). The median and mean CT-score was 1.33 (n=34) and 2.66 (n=54) in patients with and without right lung pleural effusion respectively. The same measure was 1.50 (n=34) and 3.00 (n=36) in patients with and without left lung pleural effusion respectively.

79 patients (86.8%) had CT-scores > 0 in both lungs and 12 (13.2%) patients had at least one lung with CT-score = 0.

## Discussion

From the starting day of the COVID-19 epidemic, the diagnosis of the disease became a major challenge. RT-PCR was used as the specific but not sensitive enough test. Further studies showed that the sensitivity and specificity of CT scan are totally acceptable even in comparison with RT-PCR [6-9]. This led to a dramatic increase in the number of CT scans in suspicious COVID-19 cases. Although CT scan has a remarkable role in the diagnosis of COVID-19, there is still a dilemma about its role in predicting disease severity and prognosis; so, we comprehensively studied the radiographic features of 93 COVID-19 patients to investigate the prognostic factors of chest CT scan.

Similar to other coronaviruses including SARS, the COVID-19 mostly causes constitutional symptoms including fever, cough, dyspnea, and sometimes gastrointestinal (GI) symptoms [19, 20]. Like previous studies, fever and cough are the most common symptoms in our patients with 89% and 74% prevalence respectively. However, the only symptom that was correlated with the patient’s prognosis was dyspnea. Compatible with previous studies, we found that age and underlying comorbidity (mainly pulmonary and cardiac) has a significant role in prognosis and elderly people with these comorbidities face more severe disease and worse prognosis [13, 19, 20]. Also, we found that the prognosis in men is worse than women proposing the role of gender which needs more evaluations.

Major CT scan findings in our study were consistent with previous reports [10-14]. Mixed GGO and consolidation were the most common pattern (73.7%). 47.3% was predominantly GGO and 26.4% was predominantly consolidation, followed by exclusively GGO (11%) and consolidation only (3.3%). Peripheral distribution was seen in 91.2%, of which 58% had mixed peripheral and central, bilateral involvement in 86.8% and 8.8% had normal CT scan. Compatible with previous reports, diffuse involvement of the lung was significantly more common in the poor prognosis group in our study [13, 14]. However, we also found that anterior and paracardiac involvements are related to poor prognosis. In patients with anterior lung involvement superimposed on posterior lesions and presence of density gradient we must be worried about progression to acute respiratory distress syndrome (ARDS). Although ARDS is a clinical diagnosis but classic appearance of acute ARDS in CT scan is anterior-posterior density gradient with dense consolidation present in the most dependent areas [21]. Only 9 patients (9.9%) had reactive lymphadenopathy which showed a weak impact on prognosis. Since this is a rare finding in COVID-19 patients, the determination of its role on prognosis needs more validation. We barely could differentiate pleural thickening with trace effusion so we merged these two and considered as trace effusion and found that 62.6% of patients had a trace or mild pleural effusion which was significantly more in the poor prognosis group. It was consistent with the Zhao et al study that found pleural effusion as a helpful feature in the determination of emergency type disease [14]. COVID-19 associated architectural distortion and traction bronchiectasis were proposed as poor prognostic factors [14]; nevertheless, we couldn’t find any statistically significant correlation. however, we found that the crazy-paving pattern is seen more in the poor prognosis group and could be related to the severity of the disease. On the other hand, we found that the “reversed halo sign” is more common in the good prognosis group, probably indicating the remission of the injuries. Also, the involvement of central and perivascular areas was not related with poor prognosis. These findings need further evaluations to be validated.

We calculated the CT-score for each lobe and found the mean score of each lung and the total CT-score for each patient. Since we only entered the patients with early CT scan findings (less than 8 days of symptoms onset) it is the score of disease early phase. The total CT-score was significantly higher in the poor prognosis group. The scoring methods in both previous studies were different from each other and our study [13, 14], however, they also suggested the total CT-score as a prognostic factor in COVID-19 which could help to identify the high-risk patients and give them the appropriate care. These findings imply the need to designate a single, distinct and applicable scoring system. Based on our scoring system we found that 11.5 could be an appropriate cut-off to identify the high-risk patients (sensitivity 67.4% and specificity 68.7%).We also found that the mean score of each lung is correlated with the presence of ipsilateral pleural effusion indicating that pleural effusion happens more in the severely involved lung.

Better performance of the multivariate logistic model compared to the univariate model suggests that predicting the prognosis of COVID-19 patients would be more accurate if one considers not only the total CT-score but also other CT scans, demographic and clinical findings which are reasonably expected to have a role in prognosis. We should emphasize though, the power of multivariable logistic models highly depends on the number of cases; therefore, to obtain models with more included inputs, one may need to increase the case number. The finding that the underlying comorbidity did not have a significant contribution to our model might also be the result of the same limitation.

This article faced some limitations. The number of cases was not sufficient for further analytic studies and enrollment of more factors in predicting the prognosis. We couldn’t get any follow up CT scan of the patients. All 3 hospitals were referral centers for COVID-19 patients, so it is possible that the overall CT-score of the patients in this study would not be representative of the general population. Finally, the clinical and laboratory data of the patients were not complete to be entered into the study and we couldn’t include them in multivariate analysis.

## Conclusion

Chest CT scan is a valuable imaging method in predicting the prognosis of COVID-19 disease. Among the CT findings, the crazy-paving pattern, diffuse distribution, paracardiac and anterior involvement, lymphadenopathy, main pulmonary artery dilation (above 30mm) and pleural effusion were predictors of poor prognosis, while the reversed halo sign was associated with a better prognosis. Multidisciplinary approaches consisting of clinical data, imaging features, and laboratory data would predict the prognosis more accurately.

## Data Availability

All data are available

## Acknowledgments

We thank education development office of Imam Khomeini Hospital Complex for assistance with this project and providing data.

